# A disproportionate epidemic: COVID-19 cases and deaths among essential workers in Toronto, Canada

**DOI:** 10.1101/2021.02.15.21251572

**Authors:** Amrita Rao, Huiting Ma, Gary Moloney, Jeffrey C Kwong, Peter Jüni, Beate Sander, Rafal Kustra, Stefan D Baral, Sharmistha Mishra

## Abstract

Shelter-in-place mandates and closure of non-essential businesses have been central to COVID-19 response strategies including in Toronto, Canada. Approximately half of the working population in Canada are employed in occupations that do not allow for remote work suggesting potentially limited impact of some of the strategies proposed to mitigate COVID-19 acquisition and onward transmission risks and associated morbidity and mortality. We compared per-capita rates of COVID-19 cases and deaths from January 23, 2020 to January 24, 2021, across neighborhoods in Toronto by proportion of the population working in essential services. We used person-level data on laboratory-confirmed COVID-19 community cases (N=74,477) and deaths (N=2319), and census data for neighborhood-level attributes. Cumulative per-capita rates of COVID-19 cases and deaths were 3-fold and 2.5-fold higher, respectively, in neighborhoods with the highest versus lowest concentration of essential workers. Findings suggest that the population who continued to serve the essential needs of society throughout COVID-19 shouldered a disproportionate burden of transmission and deaths. Taken together, results signal the need for active intervention strategies to complement restrictive measures to optimize both the equity and effectiveness of COVID-19 responses.

## Background

The City of Toronto, Canada’s largest city, has reported 92,644 cases and 2,619 deaths from COVID as of February 23, 2021.(1) In addition to testing and isolation, shelter-in-place mandates and the closure of non-essential businesses aiming to reduce contacts have been central to Toronto’s COVID-19 response. However, many businesses and services have been deemed essential to support the general needs of society. Specifically, Public Safety Canada defines “essential workers” as working in one of ten critical infrastructure sectors, including health, food, transportation, and manufacturing.(2) As of April 2020, an estimated 40% of the working population in Canada were essential workers employed in occupations not amenable to remote work, with the lowest likelihood of remote work among lower income households.(3) Thus, it has been suggested that shelter-in-place mandates may be insufficient at protecting essential workers from COVID-19 and associated morbidity and mortality. In this study, we evaluated the relationship between per-capita rates of COVID-19 cases and deaths and the proportion of the population working in essential services across neighborhoods in Toronto, Canada.

## Methods

We used Case and Contact Management Solutions (CCM)+ person-level data on laboratory-confirmed COVID-19 cases and deaths, and the Statistics Canada 2016 Census data for neighborhood-level attributes. The study population comprised community cases and deaths, thus excluding long-term care residents, reported between January 23, 2020 and January 24, 2021 in Toronto. Toronto has a population 2,731,571 including 51.5% who self-identify as visible minority. We stratified the city’s 3702 dissemination areas (DA) representing geographic areas including approximately 400-700 individuals into tertiles by ranking the proportion of population in each DA working in essential services (health, trades, transport, equipment, manufacturing, utilities, sales, services, agriculture). Strata 1, 2, and 3 comprised DAs across which a median of 27.8% (inter-quartile range [IQR] 23.4-31.5%), 44.7% (IQR 40.0%-50.0%), and 62.9% (IQR 58.4%-68.0%) of the population, respectively, worked in essential services. We generated per-capita daily epidemic curves using 7-days rolling averages for cases and for deaths, and cumulative per-capita rates using census-reported population of each DA. The date of report to public health for both COVID-19 cases and deaths were used for per-capita rate calculations.

## Findings

COVID-19 was initially concentrated in stratum 1 (lowest essential workers). By early April, per-capita cases were consistently higher in strata 2 and 3 (**Figure 1A**), and persisted during each period of closure of non-essential services (first lockdown March 17 to May 18, second major restriction November 23 to December 25, and a more stringent lockdown starting on December 26). By the end of the study period, cumulative rates of cases per 100,000 were 1332, 2495, and 4355 in strata 1, 2, and 3, respectively; representing a 1.9-fold and 3.3-fold higher rate than stratum 1 (**Figures 1C & D**).

**Figure 1.**
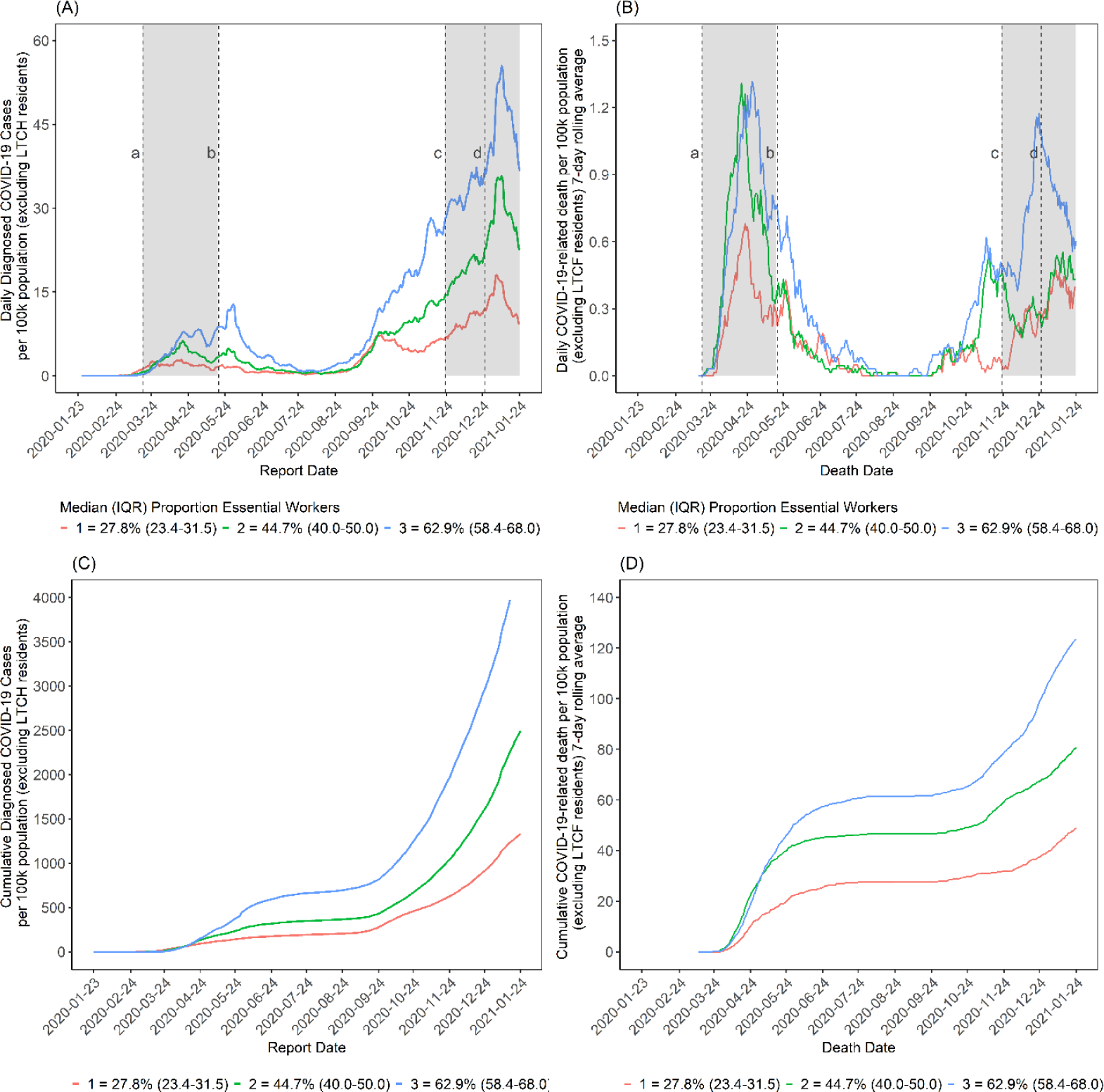
Daily per-capita COVID-19 cases (A) and deaths (B) and cumulative per-capita COVID-19 cases (C) and deaths (D) by neighbourhood-level proportion of essential workers in Toronto, Canada (January 23, 2020 to January 24, 2021). The daily per-capita rate is depicted as a 7-day rolling average. Stratum 1 represents neighbourhoods with the smallest proportion of the population working in essential services, while stratum 3 represents neighbourhoods with the highest proportion of essential workers. Cases and deaths do not include residents of long-term care homes. Essential services include: health, trades, transport, equipment, manufacturing, utilities, sales, services, and agriculture. Closure of non-essential workplaces are indicated by: (a) from the start of the first lockdown on March 17, 2020 to (b) the re-opening of the province on May 18, 2020; (c) the start of the 2^nd^-major restriction on November 23; and (d) the start of a more stringent lockdown on December 26, 2020. By the end of the study period, cumulative rates of cases per 100,000 population were 1332, 2495, and 4355 in strata 1, 2, and 3, respectively; and cumulative rates of COVID-19 deaths per 100,000 population were 49, 81, and 123 in strata 1, 2, and 3, respectively.

Per-capita COVID-19 deaths were similarly concentrated in strata 2 and 3. By the end of the study period (**Figure 1B**), cumulative rates of death per 100,000 in strata 1, 2, and 3 were 49, 81, and 123, respectively, representing a 1.9-fold and 2.5-fold higher rate than stratum 1 (**Figures 1C & D**). The number of daily per capita cases among stratum 2 was consistently between strata 1 and 3. Daily per-capita deaths fluctuated over the course of the outbreak, demonstrating similarities with stratum 3 during the first lockdown and with stratum 1 during the second major restriction.

## Discussion

In the context of shelter-in-place mandates, there have been disproportionate risks and consequences of COVID-19 borne by those living in neighborhoods with higher proportions of essential workers. The results presented here are consistent with early studies highlighting that these occupations would not be amenable to remote work, and thus people may experience sustained contact rates irrespective of restrictive measures.(3) Many of these occupations afford lower wages and are often held by people hired as contractors with unclear labor rights and generally lacking traditional employment benefits, such as paid sick leave.(4-6) Precarious financial conditions among many essential workers further limit bargaining power to demand adequate personal protective equipment and safe working conditions from employers or clients.(7, 8) Decreased beneficial impact of restrictive COVID-19 mitigation strategies is likely compounded by increased likelihood that lower income essential workers also live in multigenerational households, where an older parent or grandparent may be at a much higher risk of morbidity or mortality.(4)

The findings highlight a prevention gap with restrictive COVID-19 intervention strategies including shelter-in-place mandates. Moving forward necessitates policies and programs that actively protect workers in occupations that remain active in the context of lockdowns. Public and occupational health strategies could include primordial prevention aimed at keeping COVID-19 out of essential workplaces including through paid leave facilitating people to stay home if they have symptoms or a known exposure; primary prevention designed to limit transmission within the workplace such as onsite rapid testing and improved access to symptom assessments, and improved ventilation; secondary prevention directed at limiting the size of the outbreak including infection prevention and control measures including isolation, mass testing, and cohorting; and tertiary prevention strategies targeted at limiting outbreak-related mortality including temporary housing support to prevent transmission to households of essential workers.(9) Moreover, these data support the specific allocation of COVID-19 vaccines to particularly burdened communities with more essential workers given the intersection of higher occupational and housing risks in these areas.

In sum, these are data from a large urban center demonstrating that the communities with more essential workers who continued serving the needs of society during COVID-19 have been disproportionately affected by COVID-19 morbidity and mortality. Ultimately, these results suggest the need for active public and occupational health intervention strategies to complement restrictive measures to optimize both the equity and effectiveness of COVID-19 responses.

## Data Availability

Data used in this study can be made available upon request.

## List of Abbreviations

CCM+: Case and Contact Management Solutions
DA: dissemination areas
IQR: interquartile range

## Ethics Approval

The University of Toronto Health Sciences Research Ethics Board (protocol no. 39253) approved the study.

## Acknowledgements

Reported COVID-19 cases were obtained from the Contact Management Solutions (CCM)+ via the Ontario COVID-19 Modelling Consensus Table and Ontario Ministry of Health. We thank Kristy Yiu (MAP Centre for Urban Health Solutions, St. Michael’s Hospital) for support with data management, and the Ontario Community Health Profiles Partnership.

